# A health-system-scale, episode-resolved multimodal atlas of cancer cachexia

**DOI:** 10.1101/2025.09.29.25336906

**Authors:** Venise Jan Castillon, Amy X. Xie, Sonia Boscenco, Ethan Tse, Neil Ruthen, Jamie Wang, Ziad El Bakouny, Michele Waters, Chenlian Fu, Christopher Fong, Urvi A Shah, A Ari Hakimi, Justin Jee, Eileen White, Tobias Janowitz, Marcus D Goncalves, Ed Reznik

## Abstract

Cancer cachexia is a wasting syndrome with outsized impact on morbidity and mortality. Neither the etiology of cachexia, nor its consequences on patient physiology and outcomes, are well-understood. Here, we repurposed longitudinal clinicogenomic data from 59,493 cancer patients to define episode-resolved trajectories of cachexia and linked them to serology, tumor genotypes, and clinical outcomes. Cachexia risk concentrated around periods of disease progression and associated with inferior outcomes in nearly all cancer types. Across cancers, cachectic episodes exhibited a consistent serologic signature characterized by low albumin and hemoglobin and high levels of liver enzymes. Numerous somatic tumor genotypes, including *TP53* mutation across several diseases, were associated with elevated cachexia risk. Motivated by these observations, we developed a multivariate model to predict impending risk of cachexia in lung and colorectal cancer patients. Routine clinicogenomic data is therefore a powerful resource for discovery in cachexia and other cancer-associated pathophysiologies.

## Introduction

The uncontrolled cellular growth and local tissue disruption produced by tumorigenesis induces a complex host response, including chronic inflammation, metabolic reprogramming, and endocrine dysfunction. These systemic changes can emerge early and persist throughout the disease course, contributing to profound alterations in appetite, energy balance, tissue composition, and immune function^1,2^. Together, they underlie debilitating syndromes such as fatigue, anorexia, and cachexia. Understanding these pathophysiologies in human cancer patients requires integrated, longitudinal measurements of host physiology and tumor biology, but most human molecular studies focus narrowly on malignant tissue, overlooking the systemic consequences of cancer on the whole organism.

Cancer cachexia (herein, “cachexia”) is a paraneoplastic metabolic syndrome characterized by progressive, unintentional weight loss^1,3,4^ that believed to affect approximately half of all cancer patients^1,5^. Cachexia is associated with reduced quality of life^6,7^, premature termination of anti-cancer therapy, increased treatment complications, and elevated risk of death^8–13^. Despite its prevalence and well-acknowledged consequences for morbidity and mortality, the underlying causes of cachexia are poorly understood. Recently, murine and fly models of cancer cachexia have identified a variety of tumor-intrinsic molecular events, circulating factors, and behavioral factors that mediate cachexia and its severity^1,14–17^. However, despite these advances in model organisms, there remains a major gap in our understanding of the manifestation of cachexia, and its molecular underpinnings, in human cancer patients^18,19^. This gap exists because molecular studies of cancer largely focus (with important exceptions^20–23^) on measurements of malignant tissue, and therefore fail to capture measurements of wasting in non-malignant muscle and adipose compartments affected by cachexia.

Here, by repurposing clinicogenomic measurements collected in the routine care of 59,493 cancer patients, we interrogate the clinical evolution and molecular basis of cancer cachexia. Using this rich, longitudinally collected, multimodal dataset, we define the basic presentation and temporal evolution of cachexia across cancer types. Integrating serologic measurements of patient physiology, we discover characteristic and cancer-type-agnostic changes in blood biomarkers that define episodes of cachexia across cancer patients. Finally, through genomic analysis, we discover that certain tumor genotypes, and especially the presence of *TP53* mutations, are associated with an increased risk of cancer cachexia. We build on these discoveries to produce novel, prospective models of the impending risk of cachexia in patients from two distinct cancer types.

## Results

### Population-scale identification of cachexia in patients with cancer

To quantitatively study patterns of weight loss in patients with cancer, we obtained multimodal clinical, molecular, and radiologic data on 59,493 patients **(Fig.1A; see also Supplemental Figure 1A)**, covering 656 total distinct cancer histologies **(Supplemental Figure 1B)**, who consented to the MSK-IMPACT prospective clinical sequencing protocol as part of their routine clinical care. These data contained rich measurements of patient physiology, including longitudinal measurements of body weight, routine and specialized serologic measurements, and detailed treatment histories. The temporal granularity of the data varied significantly across modalities. For a typical patient, we had 39 (IQR 14-52) measurements of body weight per patient over an average of 60.1 (IQR 23.1-78.4) months, 51 (IQR 18-68) serological measurements per patient, 16 ascertainments of disease progression (IQR 8-21), and at least one genomic measurement.

**Figure 1.**
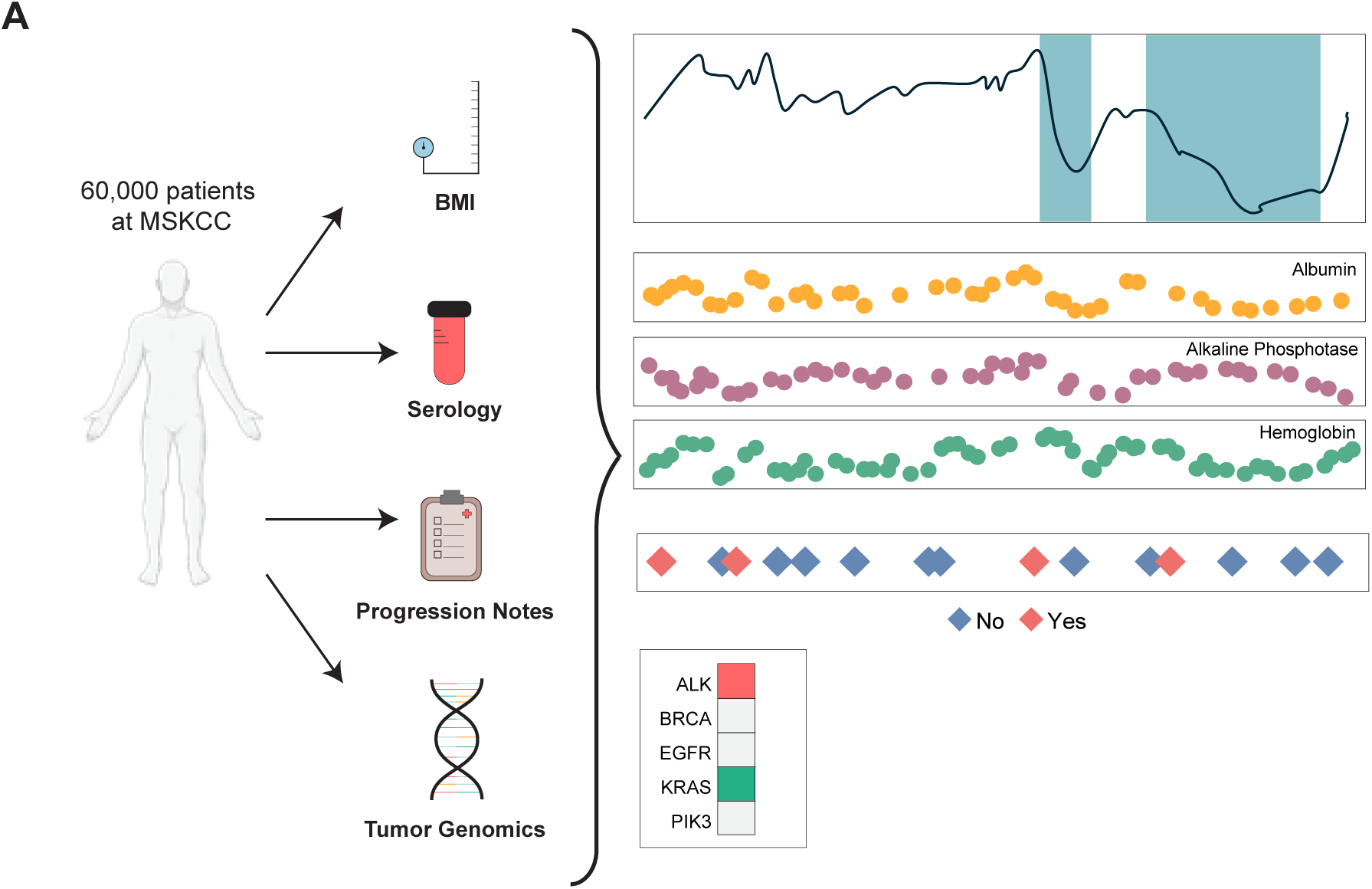
Cohort selection and multimodal data coverage. (**A**) Overview of data modalities available, including longitudinal BMI trajectories, serologic laboratory tests, progression annotations, and tumor genomic profiling.

Across this cohort, we obtained 2,335,098 individual measurements of BMI. We developed a novel framework for identifying cachexia by defining specific time intervals where a given patient’s body weight loss satisfied the clinical criteria for cachexia, *i.e.* ≥ 5% weight loss over a 6-month period (see **Methods**) **(Fig.2A; see also Supplemental Fig. 1C)**. In total, we identified 37,544 discrete episodes of cachexia across our cohort, affecting 48.8% of all patients in total **(Fig. 2B)**, with 24.1% of cachexia-positive patients experiencing more than one episode of cachexia **(Fig. 2C)**. An analogous approach was implemented to capture more severe cachectic episodes of ≥ 10% weight loss over a 6-month period (affecting 23% of all patients) and ≥ 15% weight loss over a 6-month period (affecting 9% of all patients) **(Fig 2B)**. Normalizing for total clinical follow-up, a typical patient could expect to be experiencing cachexia for 11% (IQR 0 - 13.5%) of their total follow-up time **(Supplemental Fig. 1D).**

**Figure 2.**
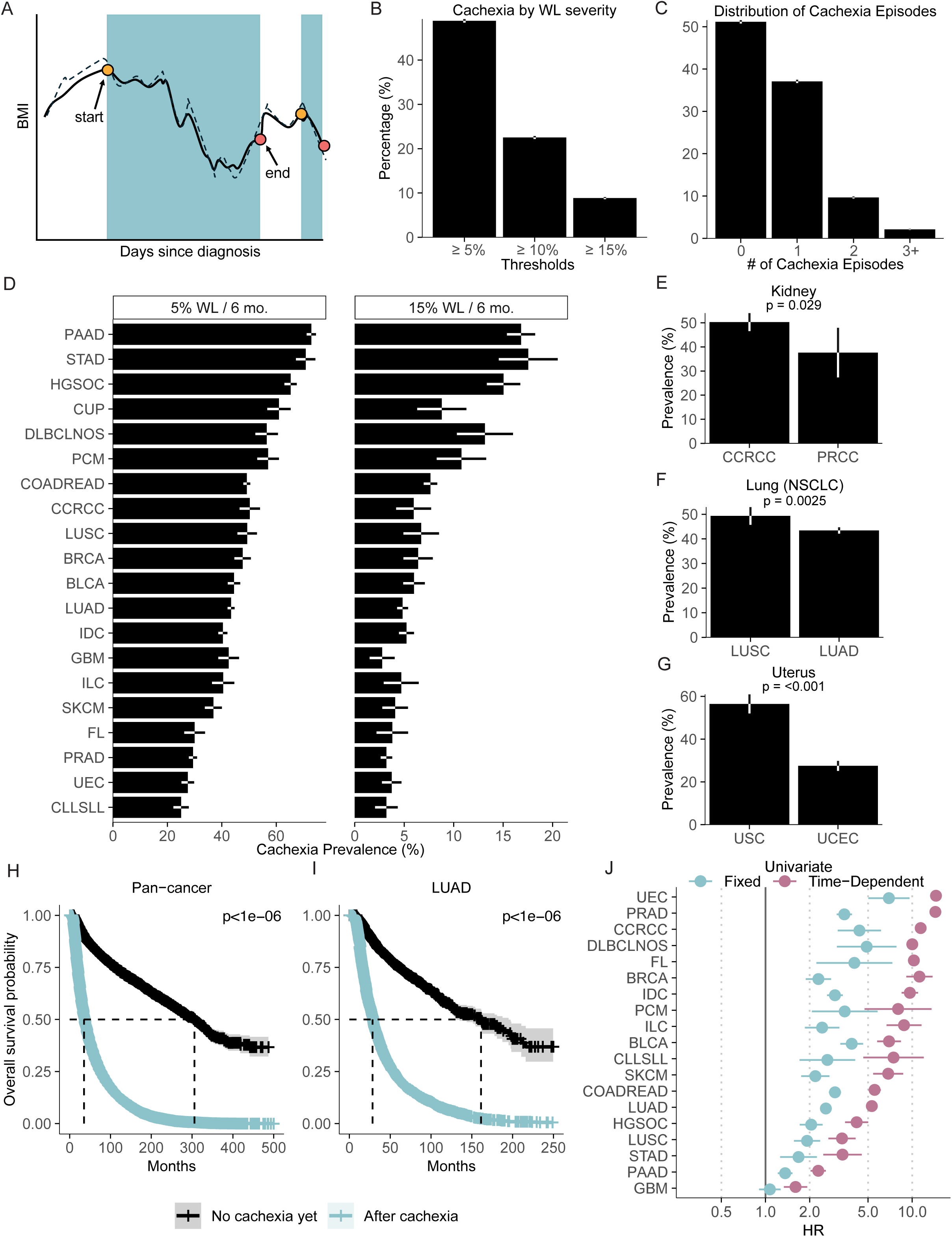
Prevalence and demographic associations of cancer cachexia. (**A**) Example BMI trajectory illustrating algorithmically defined cachexia episodes (≥5% weight loss over six months). (**B**) Prevalence of cachexia at ≥5%, ≥10%, and ≥15% weight loss thresholds. (**C**) Number of cachexia episodes per patient at the ≥5% threshold. (**D**) Cachexia prevalence by cancer type at ≥5% and ≥15% thresholds. (**E**) Cachexia prevalence in renal cancer subtypes (clear cell vs. papillary). (**F**) Cachexia prevalence in non–small cell lung cancer subtypes (adenocarcinoma vs. squamous). (**G**) Cachexia prevalence in uterine cancer subtypes (serous vs. endometrioid).(**H-I)** Overall survival of patients stratified by cachectic status either pan-cancer **(H)** or in lung adenocarcinoma **(I)**. **(J)** Association between cachexia and overall survival, treating cachexia as either a fixed or time-dependent hazard

Consistent with prior literature, the burden of cachectic episodes varied significantly across cancer types^5,24^. Several gastrointestinal malignancies showed higher prevalence of cachexia, including pancreatic adenocarcinoma (PAAD) (72.9%), stomach adenocarcinoma (STAD) (70.8%), and colorectal adenocarcinoma (COADREAD) (49.2%). Conversely, rates of cachexia were characteristically lower in those with prostate adenocarcinoma (PRAD) (29.4%), cutaneous melanoma (CM) (36.9%), and chronic lymphocytic leukemia/small lymphocytic lymphoma (CLL/SLL) (25.1%). A similar pattern of incidence was observed at the more stringent ≥15% weight loss threshold, where gastrointestinal cancers remained the most affected **(Fig. 2D)**. Rates of cachexia also varied across histologically distinct tumor types arising from the same tissue. For example, clear cell renal carcinoma patients had higher cachexia prevalence compared to papillary renal cell carcinoma patients (p□=□0.029; **Fig. 2E**). Similarly, among patients with non–small cell lung cancer, adenocarcinoma patients had a higher incidence of cachexia than squamous cell carcinoma patients (p□=□0.0025; **Fig. 2F**). Finally, in uterine cancers, the endometrioid subtype had lower prevalence than the more aggressive serous subtype (p□<□1□×□10⁻¹□; **Fig. 2G**). Similar within-tissue variation was observed in additional organs but was not universal (**Supplemental Fig. 2A**): pancreas (PAAD > PNET; p < 0.001) and blood cancers (DLBCL > FL; p < 0.001) showed significant differences, whereas liver (CHOL vs. LIHC; p = 0.76), esophagus (ESCA vs. GEJ; p = 0.74), and breast (ILC vs. IDC; p = 0.96) showed no significant difference in cachexia incidence across subtypes.

To explore differences in cancer cachexia across cancer types and clinical demographics, we carried out a regression analysis to estimate the likelihood of cachexia at tumor diagnosis across these demographic factors, considering all cancer types with >500 patients (see **Methods**). Age and sex were significantly associated with cachexia in specific cancer type contexts, including associations with age in cutaneous melanoma (log₂(OR) = −0.55, FDR = 0.018), stomach adenocarcinoma (log₂(OR) = −0.64, FDR = 0.038), and lung adenocarcinoma (log₂(OR)= −0.32, FDR = 0.0008) for patients aged ≥60, and associations with sex in lung squamous cell carcinoma (log₂(OR) = 0.54, FDR = 0.04). Notably, having higher BMI at diagnosis was associated with increased likelihood of cachexia in multiple cancers, including pancreatic adenocarcinoma (log₂(OR)= 0.83, FDR= 1.33×10^-5^), high grade-serous ovarian cancer (log₂(OR) = 0.77, FDR= 1.35×10^-4^), and renal clear cell carcinoma (log₂(OR) = 0.78, FDR=0.03) (**Supplemental Fig. 2B,** See also **Supplemental Table 1**). We then evaluated weight-loss relative to baseline BMI using the Weight-Loss Grading System (WLGS)^25^, focusing on the first cachectic episode experienced by each patient (**Supplemental Fig. 2C-G;** also see **Methods)**. Among underweight patients, nearly all episodes were severe, with 74.8% classified as WLG 4 and 24.6% as WLG 3. In contrast, obese patients rarely fell into WLG 4 (<0.2%) and instead were distributed across WLG 1 (16.3%), WLG 2 (38.0%), and WLG 3 (40.7%). This indicates that patients beginning cachexia from an underweight state almost uniformly present with high-grade weight loss later in the disease course, whereas obese patients enter cachexia across a broader spectrum of grades. Although low BMI is a hallmark of advanced cachexia, these findings suggest that patients with lower baseline BMI may be more likely to experience clinically significant weight loss and may reflect a limited physiological reserve allowing for detectable loss, or a broader phenotype of cachexia that includes patients with obesity, consistent with prior literature^8,26^.

Finally, although cachexia is widely believed to be associated with inferior patient outcomes^27,28^, quantitative data on the direct association between cachexia and patient survival is limited. We therefore carried out survival analysis using either time-independent or time-dependent Cox proportional hazards across 19 cancer types with at least 500 patients. Using time-independent/fixed-effects models, 18/19 cancer types demonstrated a significant association between cachexia and inferior outcomes with only glioblastoma patients demonstrating a lack of significant association. For example, in lung adenocarcinoma, patients who experienced cachexia had significantly reduced survival compared to those who did not (HR=2.58, FDR< 1 × 10⁻¹□; **Fig. 2H**), a pattern that was also observed at the pan-cancer level (p < 1 × 10⁻¹□; **Fig. 2I**). Extending this to time-dependent Cox models, cachexia onset was associated with inferior overall survival in 19 out of 19 cancer types **(Fig. 2J)**. The strongest effects were observed in uterine endometrioid carcinoma (HR=14.53, p< 1 × 10⁻¹□), renal clear cell carcinoma (HR=11.42, p< 1 × 10⁻¹□), and prostate adenocarcinoma (HR= 14.39, p < 1 × 10⁻¹□) where the onset of cachexia was associated with markedly increased risk of death **(Table 1)**. These results remained statistically significant even after multivariate analysis controlling for age, sex, and cancer type **(Supplemental Table 2)**. When stratified by weight-loss grade, higher WLG scores were also associated with progressively worse survival **(Supplemental Fig. 2H)**. Thus, cachexia is almost universally associated with inferior overall survival across cancer types and patient demographics.

**Table 1:**
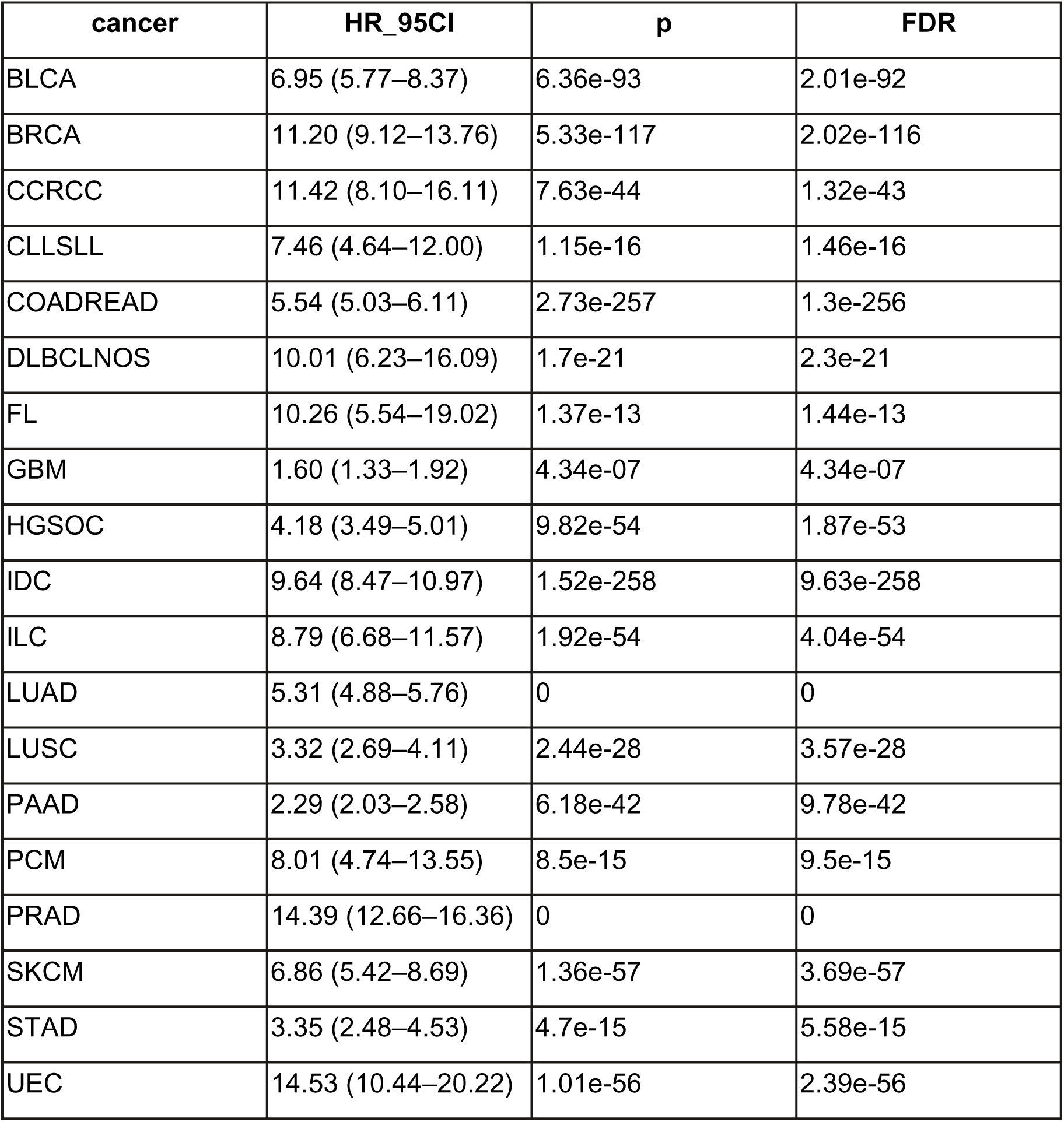
Univariate Overall Survival Time-Dependent Cox Model Results Across Cancer Type.

### Moments of disease progression are enriched during cachectic episodes

Prior studies have suggested that in most cancer types, the prevalence of cachexia is higher in those with advanced disease as opposed to early stage disease, suggesting that the most extreme forms of wasting arise late in the disease^24,29,30^. To characterize when cachexia emerges during the clinical course and its subsequent clinical evolution, we systematically mapped the temporal incidence of cachectic episodes across each patient’s clinical trajectory **(Fig. 3A)**.

**Figure 3.**
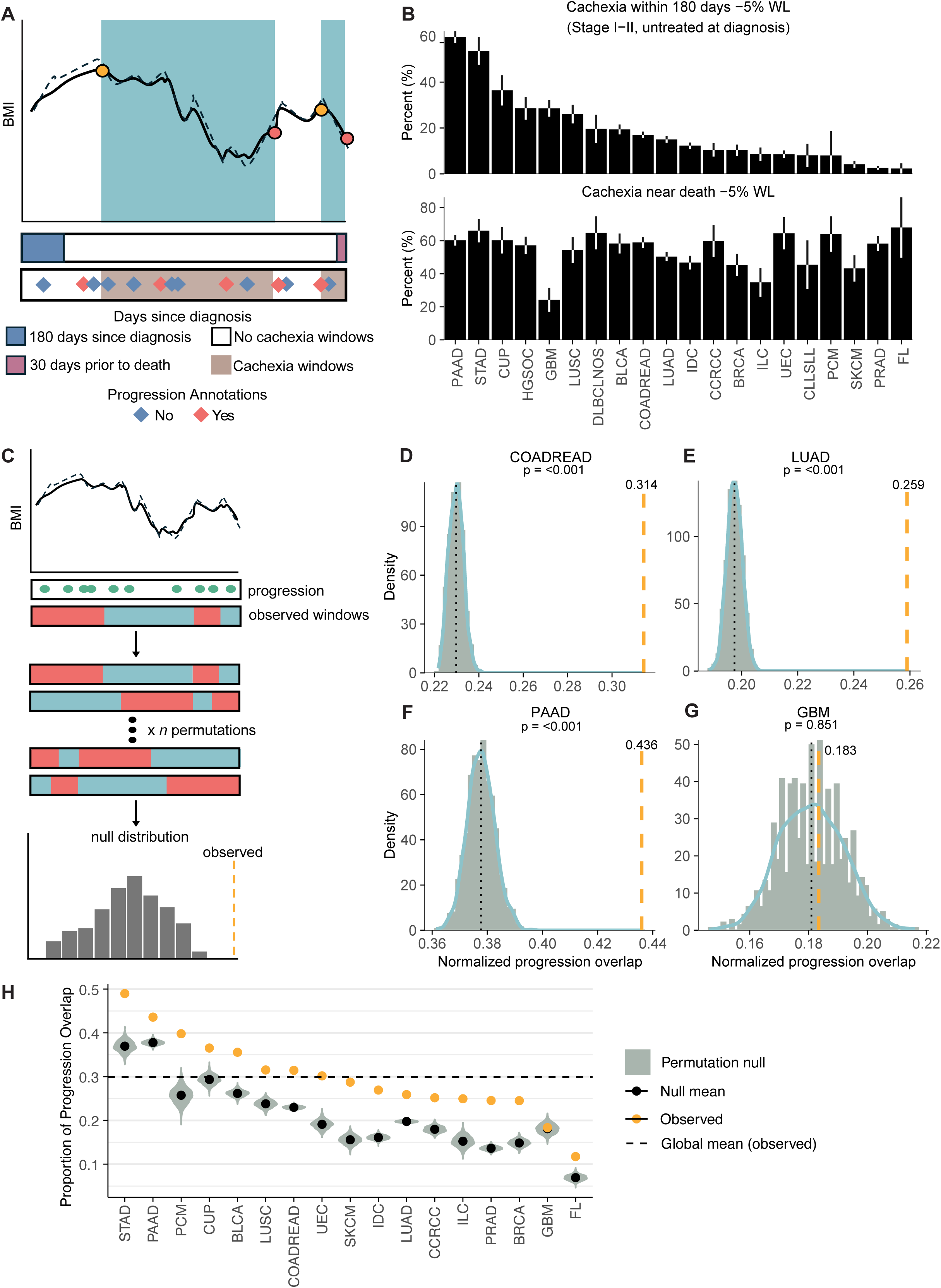
Cachexia is associated with progression on cancer therapy. **(A)** Schematic illustrating time periods surrounding diagnosis and end-of-life relative to cachexia episodes. **(B)** Cachexia within 180 days of diagnosis (Stage□I–II, untreated at diagnosis; top) and cachexia near death (±30□days; bottom) by cancer type. **(C)** Permutation framework generating null distributions of cachexia–progression overlap by random reassignment of patient spans (duration and status preserved). **(D-G)** Permutation tests for cachexia episode overlaps with progression events in **(D)** colorectal adenocarcinoma (observed exceeds null), **(E)** lung adenocarcinoma, **(F)** pancreatic adenocarcinoma, and in **(G)** glioblastoma (observed approximates null). **(H)** Summary of cachexia–progression overlap across cancer types.

Approximately 22.4% of patients with early-stage disease (stage I-II, untreated at diagnosis; n=24,151) experienced an onset of a cachectic episode in the first 180 days following cancer diagnosis **(Fig 3B)**, although this frequency dropped when considering more severe weight loss thresholds for calling cachexia (10% weight loss over 6 months: 8% of patients; 15% weight loss over 6 months: 2.7%) (**Supplemental Fig. 3A)**. Early-onset cachexia was particularly enriched in GI malignancies: relative to the other cancers, patients with pancreatic adenocarcinoma (OR=6.56, FDR < 1 × 10⁻¹□) and stomach adenocarcinoma (OR=5.2, FDR < 1 × 10⁻¹□) were more likely to develop cachexia within this early window. This enrichment remained evident at higher weight loss thresholds, with early-onset cachexia still significantly more likely in PAAD and STAD compared to other cancers (≥10% threshold: OR= 6.66, 4.2; ≥15% threshold: OR= 6.05, 3.51, respectively) **(Supplemental Table 3)**. Although lung adenocarcinoma is often considered a prototypical cachectic tumor type^31,32^, patients with LUAD in our cohort were slightly less likely to develop early-onset cachexia (OR=0.83, FDR =0.03) relative to other cancer types, with even lower odds at higher weight loss thresholds (≥10% weight loss/6 months OR = 0.6; ≥15% weight loss/6 months OR = 0.61).

Conversely, cachexia frequently emerged near the end of life, reflecting its role as a marker of advanced disease and systemic decline^1,33^. Among deceased patients with BMI follow-up in the last 30 days of life (n=11, 253), 55.1% died during or within 30 days of a cachectic episode **(Fig 3B)**. This pattern was particularly pronounced in acute myeloid leukemia (AML), esophageal adenocarcinoma (ESCA), and stomach adenocarcinoma (STAD) where patients were significantly more likely to experience cachexia near death compared to the other cancers (OR= 2.05, 1.76, and 1.67, respectively; all FDR< 0.005). Under higher weight loss thresholds, 27% of deceased patients died during or shortly after a cachectic episode at the ≥10% weight loss over 6 months threshold, and 11.3% at the ≥15% weight loss over 6 months threshold **(Supplemental Fig. 3A)**. Patients with AML, ESCA, and STAD continued to show significantly higher odds of cachexia at or near death compared to other cancers at higher weight loss thresholds (≥10%: OR=2.39, 2.24, and 1.63; ≥15%: OR = 3.22, 2.44, and 2.33, respectively; all statistically significant).

We reasoned that the elevated rate of cachexia near end-of-life may reflect an increased risk of cachexia during progression on therapy. To test this, we developed a statistical approach to determine whether progression events occurred during cachexia at a rate higher than expected by chance (see **Methods**). Using 252,099 previously-annotated progression events^34^, we performed a permutation test in which episodes of cachexia were randomly reassigned within each patient timeline, preserving the overall frequency and temporal distribution of the progression events **(Fig.3C)**. Repeating this process for n=1,000 permutations generated a null distribution of the expected overlap between instantaneous moments of disease progression and time-delimited cachectic episodes. Overall, 16 of 17 cancer types with at least 500 patients showed a significant enrichment in overlap between disease progression and cachectic episode **(Fig. 3H)**. For example, in colorectal adenocarcinoma, nearly one-third of progression events (observed overlap = 0.31) occurred during cachexia episodes, compared to an expected fraction of 0.23 under random permutation (p < 0.001; **Fig. 3D**). A similar enrichment was observed in lung adenocarcinoma (observed = 0.26, null = 0.20, p < 0.001; **Fig. 3E**) and pancreatic adenocarcinoma (observed = 0.44, null = 0.38, p < 0.001; **Fig. 3F**). By contrast, glioblastoma multiforme showed no significant deviation from expectation (observed= 0.183, null = 0.181, p = 0.819; **Fig. 3G)**. These findings were also observed under a more stringent ≥15% weight-loss threshold **(Supplemental Fig. 3B)**, indicating that the enrichment persists with severe weight loss.

### Disease-agnostic serologic signatures reveal systemic perturbations in cachexia

Beyond weight loss, cachexia disrupts systemic physiology, leading to widespread metabolic and inflammatory perturbations^35–38^. This systemic involvement is recognized across other chronic conditions^39^, including cancer, where persistent inflammation and tumor-derived factors further exacerbate catabolic processes^40–42^. Interrogating such systemic phenotypes is challenging with tumor-tissue-focused molecular data such as DNA or RNA sequencing, but highly amenable to routinely collected blood-lab measurements of physiology from complete blood counts and comprehensive metabolic panels.

To assess the physiologic impact of cachexia, we examined 30 routinely collected longitudinal serologic markers, comparing trends between cachectic and non-cachectic time periods (see Methods) **(Fig. 4A)**. To control for patient-intrinsic heterogeneity, we utilized mixed effects models that modeled patient-specific trends in lab values while adjusting for sex as well as longitudinal sampling frequency, directly comparing lab values during cachectic weight loss to lab values outside of cachectic weight loss. The association between cachexia and each serologic marker was highly consistent across cancer types, suggesting the presence of a disease-agnostic signature of cachectic weight loss. Nutritional markers such as albumin showed the strongest decreases during cachexia (mean log₂(OR) = –0.95; significant in 14 of 15 cancer types), alongside reductions in total protein (–0.50; 13 of 15), indicating pronounced nutritional depletion. Several hematologic markers were also consistently lower, including hemoglobin (–1.13; 15 of 15), hematocrit (–1.07; 15 of 15), and red blood cell count (–0.90; 15 of 15), consistent with anemia and impaired erythropoiesis. In contrast, inflammatory and tissue stress markers were elevated, including white blood cells (WBC; +0.01; 10 of 15), alkaline phosphatase (ALK; +0.55; 15 of 15), and neutrophils (+0.35; 15 of 15) **(Fig. 4B, Supplemental Table 4)**. When aggregated across diseases **(Fig. 4C)**, these directionally consistent shifts converged on a tissue!Zagnostic signature, highlighting a physiologic phenotype of cachexia marked by nutritional depletion, anemia, hepatic stress, and systemic inflammation.

**Figure 4.**
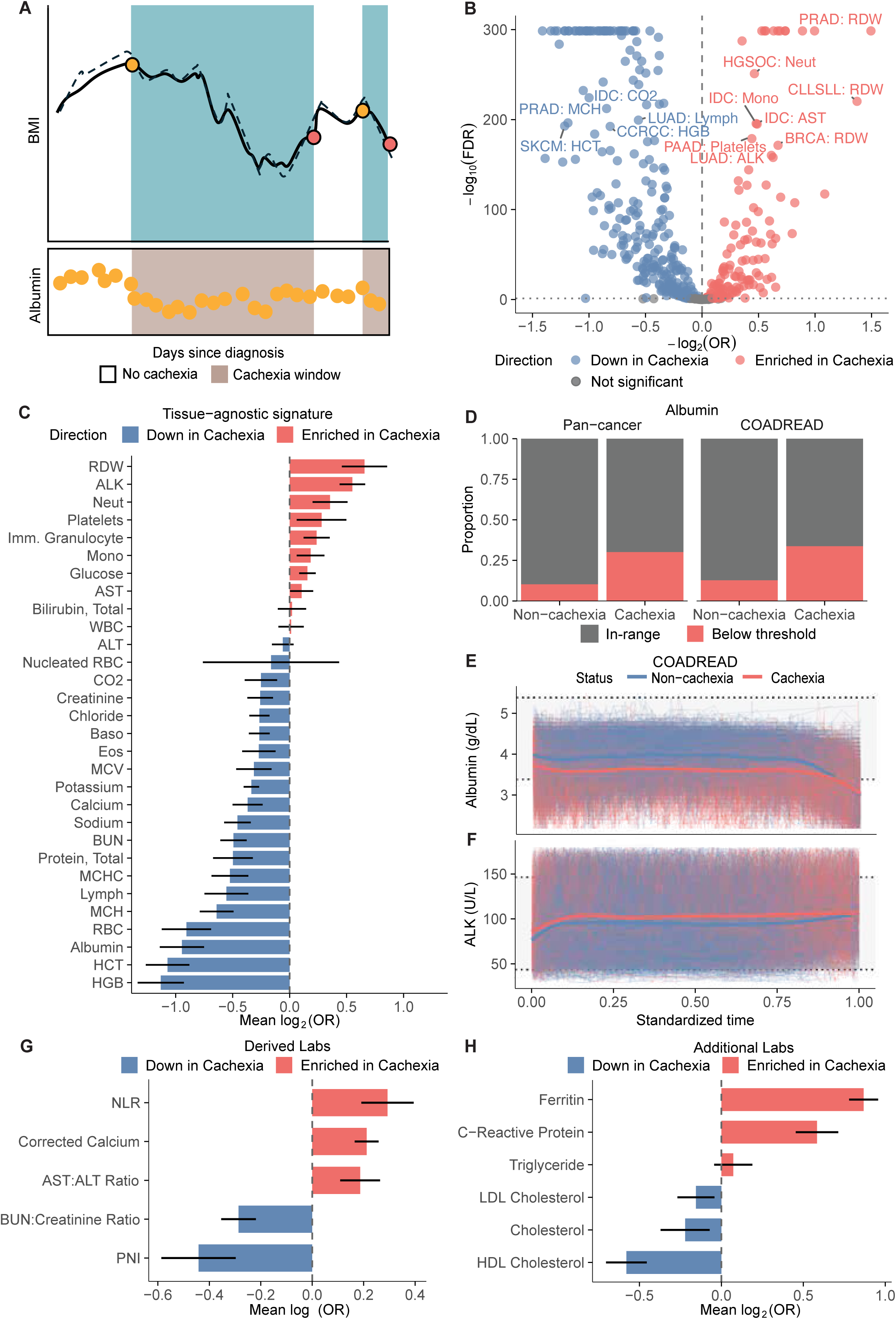
A serologic signature of cancer cachexia. **(A)** Schematic depicting measurements of albumin overlaid with a BMI trajectory. To identify signatures of cachexia, lab values in the shaded region are compared to those in the non-shaded region. **(B)** Results of univariate analysis identifying differences in lab values during cachectic episodes relative to lab values outside of cachectic episodes. **(C)** Tissue-agnostic summary of lab value associations with cachexia, derived from mixed-effects models. **(D)** Comparison of albumin values within and outside of cachectic episodes in colorectal adenocarcinoma patients (left) and pan-cancer (right). **(E)** Albumin levels are consistently lower, across all relevant time points, during cachectic episodes in colorectal cancer. **(F)** ALK levels are consistently elevated during cachectic episodes in colorectal cancer, with especially notable elevation near the end of cachexia. **(G)** Derived laboratory values, such as NLR and PNI, are systematically altered in cachectic time intervals relative to non-cachectic time intervals. **(H)** Non-CBC/CMP lab values reflecting inflammatory or nutritional status are altered in cachectic time intervals relative to non-cachectic time intervals.

Albumin, a key indicator of nutritional status and inflammation^43–45^, showed one of the strongest decreases. Most albumin values, however, still remained within the normal reference range (3.4–5.4□g/dL), indicating that cachexia often drove subtle declines rather than overt hypoalbuminemia. Across all cancer types, 30% of albumin measurements during cachectic episodes fell below the normal range, compared to 10.2% during non!Zcachectic intervals **(Fig. 4D)**. In colorectal adenocarcinoma (COADREAD), 33.7% of albumin values during cachectic windows were below 3.4□g/dL compared to 12.8% in non!Zcachectic windows, corresponding to 3.5-fold increased odds (p□<□1□×□10⁻□) of subnormal albumin during cachexia **(Fig. 4D)**. These findings indicated that while cachexia often did not produce overt hypoalbuminemia, it still drove a clinically meaningful downward shift in albumin levels. In COADREAD, for example, albumin declined more steeply during cachectic spans than during non!Zcachectic periods (slope difference□=□0.10, p□=□3.6□×□10⁻□), despite both trajectories largely remaining within the normal range **(Fig. 4E)**. In contrast to the declines observed in albumin, ALK, a marker of hepatic and skeletal stress^46^, showed more frequent elevations above the reference range (44 to 147□IU/L) during cachexia. In COADREAD, 42.2% of ALK measurements during cachectic windows exceeded the upper limit compared to 24.4% during non!Zcachectic periods, a pattern also observed across cancer types, where 24.9% of ALK values were elevated compared to 13.1% outside cachexia **(Supplemental Fig. 3A)**. ALK levels also increased temporally during cachectic spans (slope difference□=□26.21, p□=□3.1□×□10⁻□), reflecting a distinct metabolic and organ!Zsystem response to cachexia **(Fig. 4F)**.

To capture broader systemic changes beyond individual lab values, we derived five composite indices from routine clinical labs (see **Methods**), focusing on markers of inflammation and nutritional status. The neutrophil-to-lymphocyte ratio (NLR), a marker of systemic inflammation^1,47^, was significantly elevated during cachexia in 14 of 15 cancer types (mean log (OR) =0.29). The prognostic nutritional index (PNI), calculated from albumin and lymphocyte count, was significantly decreased (−0.442; 15 of 15 cancer types), indicating nutritional depletion during cachectic windows. Other indices showed consistent shifts across cancer types, including an increase in the AST/ALT ratio in 11 of 15 cancer types (+0.19; 95% CI: 0.11-0.26), an increase in corrected calcium (+0.21;0.17-0.26, in 15 of 15 diseases), and a decrease in the BUN/creatinine ratio (−0.29) in 15 of 15 cancer types, further supporting the presence of systemic physiologic alterations during cachexia **(Fig 4G)**.

Finally, we examined six additional serologic markers that, while not routinely ordered, were available in a subset of patients. Among these, C-reactive protein (CRP), a direct readout of IL-6–mediated inflammation^43,48^, was significantly elevated during cachexia in 15 of 27 cancer types (mean log₂(OR) =0.58; 95% CI= 0.45-0.71), with the strongest associations observed in pancreatic adenocarcinoma (OR =2.37, FDR = 1.1 × 10⁻□) and lung adenocarcinoma (OR = 2.18, FDR < 1 × 10⁻¹□). Although the overall proportion of abnormal CRP values, defined by the clinical reference range of 0–10 mg/L, was modest (17.8% in cachectic vs. 7.8% in non-cachectic windows), elevated CRP was significantly more frequent during cachectic periods (OR = 2.56, 95% CI: 2.42–2.71; p < 1 × 10⁻¹□) **(Fig 4H).** This difference in abnormal values suggests that while CRP elevations are statistically associated with cachexia, the clinical signal may be subtle and potentially confounded by tumor burden.

### Somatic mutations to TP53 and other genes are associated with an elevated risk of cachexia

Somatic driver mutations are determinants of therapeutic vulnerability, metastatic competency, and clinical outcome. Recent evidence suggests that certain somatic mutations, such as *STK11* loss-of-function mutations in lung adenocarcinoma, may also directly promote cancer cachexia^49–51^. However, testing the hypothesis (at population-scale) that somatic mutations may mediate risk of cachexia requires granular information on the timing of cachexia as well as rich clinicogenomic data.

We used competing risk models to calculate the cumulative incidence of cachexia over time since tumor diagnosis, while properly accounting for death as a competing event, testing all cancer-type-gene combinations with at least 5% mutant cases in a given disease in our cohort (see **Methods**). We identified 35 significant associations (FDR-adjusted p-value < 0.1) linking 22 somatic mutations to cachexia risk across 13 distinct cancer types (**Table 2**). In colorectal adenocarcinoma, mutations of three genes, *TP53* (HR= 1.17, FDR = 0.01), *SMAD4* (HR= 1.34, FDR = 9 × 10^-6^) and *KRAS* (HR= 1.16, FDR = 2 × 10^-3^) were significantly associated with higher cumulative incidence of cachexia, whereas mutations of 6 genes (*ARID1A, KMT2B, PTEN, SOX9, TCF7L2, FBXW7*) were significantly associated with lower cumulative incidence of cachexia in this disease. Notably, mutations in *TP53* were significantly associated with an increased risk of cancer cachexia in 9 cancer types, including lung adenocarcinoma **(Fig. 5C)**, breast invasive ductal carcinoma, and prostate adenocarcinoma. Conversely, several mutations—such as those in *KMT2B* and *ARID1A*—were associated with a significantly lower risk of cachexia in multiple cancer types, including colorectal adenocarcinoma **(Fig. 5D)**, suggesting a potential protective effect.

**Figure 5.**
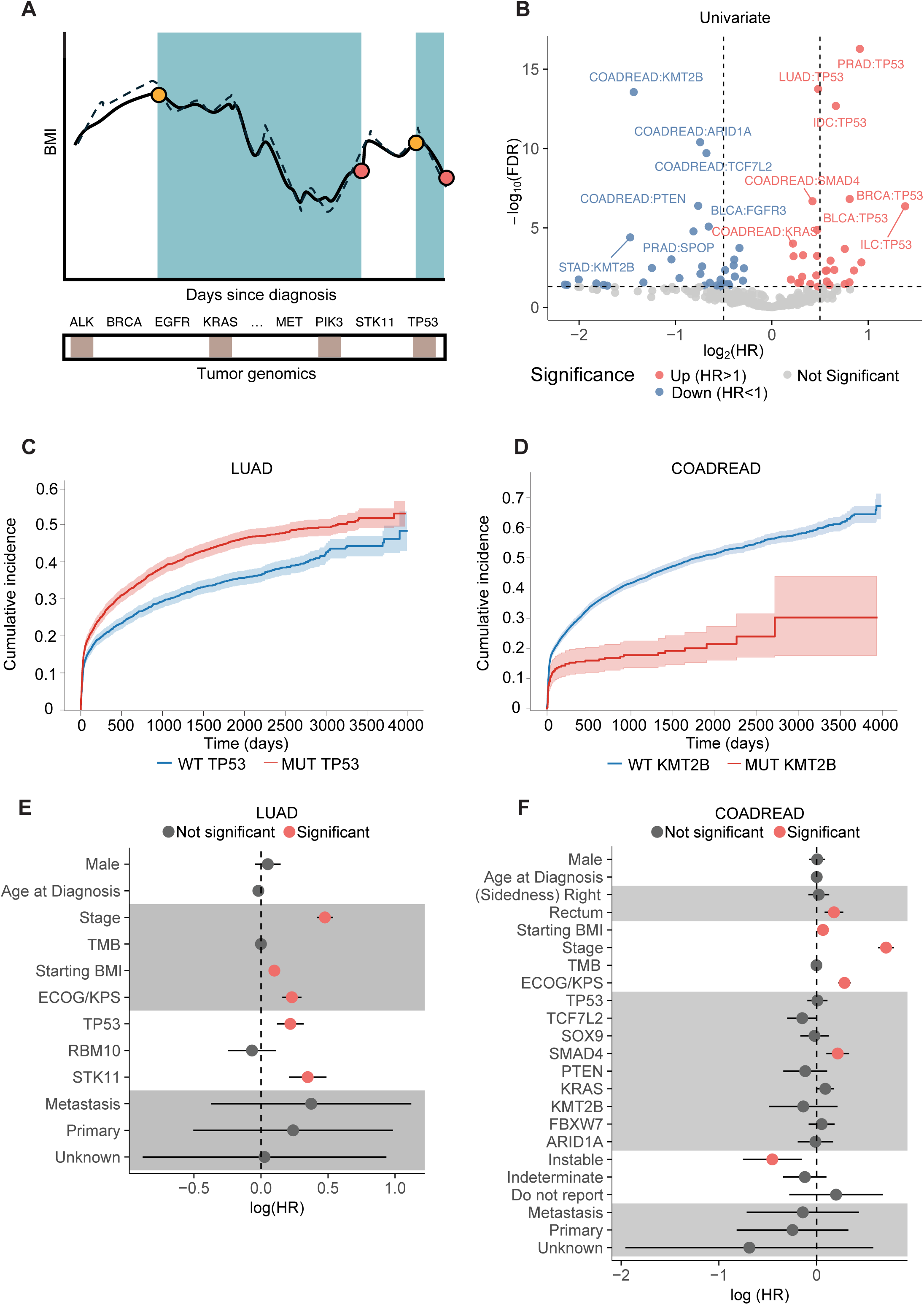
Driver mutations are associated with risk of cachexia. **(A)** Schematic **(B)** Univariate association between the presence of driver mutations and risk of cachexia from a competing Cox regression. **(C)** *TP53* mutations are associated with increased risk of cachexia in lung adenocarcinoma. **(D)** *KMT2B* mutations are associated with decreased risk of cachexia in colorectal adenocarcinoma. **(E-F)** Multimodal risk models for cachexia in lung adenocarcinoma **(E)** and colorectal adenocarcinoma **(F)**.

**Table 2:** Univariate Competing-Risks Regression Results Across Cancer Type-Gene Pairs.

| cancer | mutation | HR | p | FDR |
| --- | --- | --- | --- | --- |
| STAD | KMT2B | 0.36 | 3.96e-05 | 9.70e-04 |
| COADREAD | KMT2B | 0.37 | 2.79e-14 | 3.19e-12 |
| ILC | TP53 | 2.61 | 4.33e-07 | 1.49e-05 |
| UEC | POLE | 0.42 | 3.36e-03 | 4.26e-02 |
| UCEC | ARID1A | 0.49 | 9.59e-04 | 1.53e-02 |
| UCEC | TP53 | 1.91 | 1.47e-03 | 2.10e-02 |
| PRAD | TP53 | 1.88 | 5.28e-17 | 1.81e-14 |
| CCRCC | MTOR | 1.81 | 4.80e-03 | 5.14e-02 |
| PRAD | SPOP | 0.57 | 1.65e-05 | 4.36e-04 |
| BRCA | TP53 | 1.75 | 1.51e-07 | 7.42e-06 |
| COADREAD | PTEN | 0.59 | 4.06e-07 | 1.49e-05 |
| UEC | FGFR2 | 1.69 | 2.09e-04 | 4.22e-03 |
| COADREAD | ARID1A | 0.6 | 4.00e-11 | 2.74e-09 |
| UTUC | FGFR3 | 0.6 | 7.85e-03 | 7.92e-02 |
| SKCM | TP53 | 0.61 | 2.66e-03 | 3.51e-02 |
| COADREAD | TCF7L2 | 0.62 | 1.94e-10 | 1.11e-08 |
| IDC | TP53 | 1.59 | 2.08e-13 | 1.78e-11 |
| BLCA | FGFR3 | 0.63 | 8.24e-06 | 2.57e-04 |
| CCRCC | BAP1 | 1.57 | 4.40e-03 | 5.14e-02 |
| UEC | TP53 | 1.53 | 1.15e-03 | 1.71e-02 |
| PRAD | APC | 1.49 | 4.77e-03 | 5.14e-02 |
| UEC | JAK1 | 1.47 | 5.04e-03 | 5.24e-02 |
| UEC | PTEN | 0.71 | 4.52e-03 | 5.14e-02 |
| LUAD | TP53 | 1.4 | 1.80e-14 | 3.08e-12 |
| BLCA | TP53 | 1.39 | 1.34e-05 | 3.84e-04 |
| BLCA | RB1 | 1.38 | 5.78e-04 | 1.04e-02 |
| SKCM | BRAF | 1.38 | 9.79e-03 | 9.59e-02 |
| COADREAD | SMAD4 | 1.34 | 2.08e-07 | 8.90e-06 |
| IDC | GATA3 | 0.76 | 2.29e-03 | 3.14e-02 |
| LUAD | RBM10 | 0.76 | 9.78e-04 | 1.53e-02 |
| COADREAD | FBXW7 | 0.79 | 1.84e-04 | 3.95e-03 |
| LUAD | STK11 | 1.25 | 5.23e-04 | 9.96e-03 |
| COADREAD | SOX9 | 0.82 | 3.48e-03 | 4.26e-02 |
| COADREAD | TP53 | 1.17 | 6.09e-04 | 1.04e-02 |
| COADREAD | KRAS | 1.16 | 9.68e-05 | 2.21e-03 |

To further characterize the robustness of genotype–cachexia associations across a spectrum of cachexia severity, we conducted additional univariate analyses using more stringent weight loss thresholds to define cachexia episodes. When applying a >10% weight loss criterion, 27 out of the original 35 gene–cancer-type associations remained statistically significant (FDR<0.1) (**Supplemental Fig. 4A, Supplemental Table 5**), indicating consistent associations with more severe manifestations of cachexia. Further increasing the threshold to >15% weight loss, 12 associations retained significance (FDR<0.1) (**Supplemental Fig. 4B, Supplemental Table 6**), demonstrating the persistence of certain genotype-risk relationships even in the context of advanced cachexia. Notably, the association of *TP53* mutations with increased cachexia risk remained significant in lung adenocarcinoma and breast invasive ductal carcinoma across all severity levels. Similarly, protective associations involving *KMT2B* and *ARID1A* mutations in colorectal adenocarcinoma were preserved at the higher thresholds. The persistence of these associations at higher severity thresholds highlights a subset of somatic mutations that are consistently correlated with cachexia burden. To further corroborate the robustness of our findings, we performed a post-hoc validation by restricting the analysis to clonal mutations— those likely to be early, ubiquitous events in tumor evolution. We re-ran the univariate competing risks regression using only clonal mutation calls for the previously identified significant gene–cancer type pairs. In total, 33 out of the 35 associations remained statistically significant, confirming the robustness of our findings (**Supplemental Table 7)**.

Associations between tumor genotypes and cachexia risk identified in univariate analysis can still be confounded by demographic factors and other clinical covariates. To address this, we performed a post-hoc multivariate analysis to adjust for these clinical variables and isolate the independent effect of oncogenic mutations on cachexia risk. We controlled for age at diagnosis, sex, sample type, TMB (Tumor Mutational Burden), microsatellite instability (MSI) status, ECOG-KPS score (Eastern Cooperative Oncology Group and Karnofsky Performance Status), cancer stage and baseline BMI. Multivariate analysis confirmed tumor genotype-cachexia associations independent of other clinical factors in 15 out of 35 cancer-type-gene associations that were found significant in univariate tests. *TP53* (HR=1.24, FDR=3.7 × 10^-5^) and *STK11* (HR=1.42, FDR=4.6 × 10^-6^) were associated with increased cachexia risk independent of other clinical factors in lung adenocarcinoma (**Figure 5E, Supplemental Table 8**). *SMAD4* (HR=1.24, FDR=1.3 × 10^-3^) was associated with increased cachexia risk independent of other clinical factors in colorectal adenocarcinoma (**Figure 5F, Supplemental Table 9**). These analyses confirm that the increased risk of cachexia in patients with *TP53* and other mutations cannot be explained by the presence of more advanced disease. We conclude from these analyses that certain tumor-intrinsic genotypes are associated with a differential risk of developing cancer cachexia.

Motivated by the broad application of existing risk models in oncology to predict patients’ prognosis and risk of recurrence, we developed multivariate predictive models to estimate the impending risk of cancer cachexia in two diseases, lung adenocarcinoma and colorectal carcinoma. We selected covariates that were significantly associated with cachexia risk from the previous multivariable regression analysis as the inputs to our model. This included clinical cancer stage, ECOG-KPS performance status, *TP53, STK11* mutation status, and baseline BMI in the lung adenocarcinoma model. In the colorectal carcinoma model, we included clinical cancer stage, ECOG-KPS performance status, *SMAD4* mutation status, microsatellite instability status, tumor sidedness and baseline BMI. These variables together were used to compute individualized risk scores for cachexia over time. We evaluated the models using bootstrapped sampling across 200 iterations. The average concordance index (C-index) was 0.642 (95% CI, 0.631–0.655) in the colorectal carcinoma model and 0.649 (95% CI, 0.636–0.662) in the lung adenocarcinoma model, indicating moderate discriminative ability. Models trained on randomly selected subsets of 50% of the cohort and tested on the remaining 50% of patients demonstrated good calibration between predicted and observed 1-, 2-, and 3-year cachexia incidence in lung adenocarcinoma (**Supplemental Fig. 4C**) and colorectal adenocarcinoma (**Supplemental Fig. 4E**). Time-dependent ROC analysis showed AUCs of 0.666, 0.711, and 0.727 for the cachexia risk prediction at 1, 2, and 3 years, respectively, in lung adenocarcinoma (**Supplemental Fig. 4D**), and AUCs of 0.645, 0.688, and 0.709 for the cachexia risk prediction at 1, 2, and 3 years in colorectal adenocarcinoma (**Supplemental Fig. 4F**). These results corroborate the accuracy of our models and support the potential clinical utility of our multivariate predictive model in early identification of high-risk patients and the development of proactive interventions.

## Discussion

Previous human studies of cancer cachexia provided prevalence estimates for select cancers and some candidate biomarkers but lacked power to resolve temporal dynamics or genomic determinants^23,24^. In contrast, this analysis of 59,493 patients and >2 million BMI measurements demonstrates that cachexia arises in discrete episodes, is enriched around progression, and is associated with robust serologic phenotypes and somatic tumor genotypes.

Cachexia affected nearly half of all patients in our cohort, presenting with diverse timing patterns across cancer types. While some malignancies, such as pancreatic and gastric adenocarcinomas, showed early onset of cachexia, others exhibited episodes later in the disease course or near treatment resistance. These episodes frequently recurred and often clustered around transitions in therapy or clinical decline, highlighting cachexia as a dynamic phenotype rather than a static event. Physiologically, cachexia was accompanied by conserved serologic signatures across cancers, including reductions in albumin, hemoglobin, and red blood cell counts, consistent with nutritional depletion and anemia, and elevations in inflammatory and tissue stress markers such as alkaline phosphatase, white blood cells, and neutrophils. These changes support a view of cachexia as a systemic metabolic and inflammatory state, where tumor burden and host response converge to drive catabolism and tissue loss.

Consistent with longstanding clinical observations^3,52^, gastrointestinal malignancies emerged as the cancers most strongly associated with cachexia in our cohort. Patients with pancreatic and gastric adenocarcinomas exhibited the highest prevalence of early-onset and recurrent cachectic episodes, a finding that is not unexpected given the central role of the gastrointestinal tract in nutrient assimilation and energy balance. Multiple mechanisms likely converge in these diseases to amplify cachexia risk: tumor-induced obstruction and altered motility impair oral intake, while mucosal disruption and surgical resections compromise nutrient absorption^53^. In addition, the dense inflammatory microenvironments of pancreatic and gastric tumors promote systemic cytokine release that suppresses appetite and drives catabolism^54^. These mechanisms are further compounded by chemotherapy^55,56^, which in gastrointestinal cancers often involves intensive cytotoxic regimens that directly damage the intestinal epithelium, exacerbate nausea and anorexia, and accelerate the onset of malnutrition^57^. Together, these pathways explain why gastrointestinal tumors consistently dominate the landscape of cancer cachexia and reinforce the urgent need for interventions that preserve appetite, nutrient absorption, and host metabolism in this high-risk group.

Tumor-intrinsic molecular features were also associated with cachexia risk. *TP53* mutations were consistently linked to increased cachexia risk across multiple cancers, while other alterations, such as *ARID1A* and *KMT2B*, were associated with lower risk in specific diseases, suggesting that distinct oncogenic programs modulate host systemic physiology. These associations point toward a biological interplay in which tumor genotype may influence inflammatory signaling, metabolic reprogramming, or tissue cross-talk driving cachexia. Building on these findings, we integrated molecular, clinical, and physiologic variables into a predictive risk model capable of stratifying patients by their likelihood of developing cachexia. This model, while moderate in predictive power, highlights that cachexia risk can be quantified early and suggests a path toward incorporating cachexia surveillance into oncology practice, enabling proactive interventions such as nutritional, metabolic, or rehabilitative support.

A major open question in the cachexia field, which this study does not address, is the etiologic cause of weight loss. While our analyses revealed strikingly consistent physiologic and serologic signatures of cachexia across diverse cancer types, it is unlikely that this syndrome arises from a single causative factor. Weight loss in cancer can reflect a spectrum of mechanisms, including structural barriers to feeding, altered appetite and behavior^58,59^, systemic inflammation^42,43,49^, and cytokine signaling^14^, and direct metabolic effects of tumors or therapy^25,57,60^. The challenge for the field is to disentangle these overlapping drivers, as no single pathway is likely sufficient to explain the breadth of cachexia observed in patients. Our framework of defining cachectic time intervals provides a path toward subtyping by aligning molecular, physiologic, and radiographic features with discrete episodes of wasting. Identifying such subtypes could ultimately enable rational, precision approaches to cachexia management tailored to the dominant mechanisms at play in each patient. This would be, in other words, a precision medicine approach to cancer cachexia.

Cachexia, like many paraneoplastic syndromes, is a physiologic phenomenon quantitatively captured by simple measurements taken during routine clinical care. Many of the key discoveries reported here, ranging from the incidence of cachexia to its characteristic serologic signature, are the direct consequence of careful analysis of otherwise routine data. While the major focus of contemporary cancer biology is on the molecular and cellular factors driving tumorigenesis, disease progression, and therapeutic response, our approach here motivates the alternative idea that clinical data itself has the potential to address major questions in cancer biology. Such data comes at no cost and at a scale that dwarfs molecular data, even when the collection of molecular data is clinically indicated. The space of clinically significant questions that could be addressed with a similar approach is likely very large.

### Limitations

Our study has several key limitations. The most significant of these limitations is that weight loss is a complex convolution of loss of different tissue compartments, including but not limited to the density and area of muscle and fat. The key challenge of future work will be refining the observations made here with detailed measurements of body composition. We acknowledge that therapeutic exposure likely modifies the risk of developing cachexia and its subsequent consequences; sophisticated statistical models will be necessary to adequately capture the convolved effects of different combinations of therapy on cachexia, and of cachexia on therapeutic receipt and efficacy. In addition, the consensus definition of cachexia specifies weight loss that is unresponsive to conventional nutritional support, yet we were unable to confirm whether patients were challenged with such support. Our single-center design within a tertiary cancer hospital may further limit generalizability to community or non-tertiary settings. Finally, as with all observational analyses, our findings establish associations rather than causation, underscoring the need for prospective interventional studies to validate the mechanisms and biomarkers identified here.

## Methods

### Patient Cohort

The cohort was derived from MSK-IMPACT, a prospective observational study of tumor evolution at Memorial Sloan Kettering Cancer Center. We obtained multimodal clinical, genomic, and serologic data on 79,240 patients consented to the MSK-IMPACT prospective clinical sequencing protocol as part of their routine clinical care. Patients provided written consent for use of their genomic data for research. The dataset also included clinical data including longitudinal BMI, treatment details, longitudinal serological tests such as complete blood panel and complete metabolic panel. We focused our analysis on 59,493 patients with a minimum clinical follow-up of 180 days following diagnosis and with at least 2 body mass index (BMI) measurements.

### Cachexia Identification

Cachectic episodes were identified using weight loss criteria consistent with established clinical definitions, primarily a ≥5% body weight loss within a 6-month window^8^. To simplify the detection of cachectic episodes, we simplified one aspect of the Fearon criteria: we applied the same body weight loss threshold to patients regardless of BMI, rather than (as in the Fearon criteria) applying a different threshold to patients with BMI < 20. To robustly capture these episodes from longitudinal body mass index (BMI) measurements, patient BMI trajectories were first smoothed using an exponentially weighted moving average (EWMA) with a smoothing factor (α) of 0.2. This reduced short-term fluctuations while preserving longer-term trends in weight loss. Following smoothing, a sliding window approach was used to detect each patient’s BMI trajectory for episodes meeting the cachexia threshold. Specifically, a drop in BMI equivalent to ≥5% weight loss relative to a preceding value within a 180-day period triggered the start of a potential cachectic window. Overlapping windows were merged into single episodes to account for prolonged periods of weight loss, and only those with a minimum duration of 15 days and a cumulative BMI loss of at least 5% were retained. Additional cachexia labels were generated for sensitivity analyses using more stringent thresholds of ≥10% and ≥15% weight loss. These episodes underwent the same detection, merging, duration, and QC criteria as the 5% threshold.

Episodes were also subjected to edema-based quality control. Instances of rapid weight fluctuation, specifically, a ≥5% weight loss followed by a ≥5% gain within a 30-day window were flagged as potentially confounded by fluid shifts rather than true tissue loss. These episodes were excluded from the final cachexia dataset.

Some clinical criteria for cachexia apply separate weight loss thresholds for patients of different BMI categories (e.g. >2% weight loss over a 6-month period for patients under a BMI of 20). Such criteria are challenging to implement in our algorithmic approach (for example, because patients may begin at BMI > 20 and subsequently cross the threshold of BMI=20 during a weight loss episode), and we leave the implementation of such refined criteria for future work.

### Data Preprocessing

#### Serological Lab Values

We collected 30 routinely collected serologic markers from complete blood count (CBC) and comprehensive metabolic panel (CMP) tests. All measurements were converted to standard clinical units to ensure consistency across patients and timepoints. To minimize noise, we excluded biologically implausible values and restricted analysis to measurements collected during active clinical follow-up.

In addition to individual serologic measurements, we computed five derived indices from routinely collected lab tests to better capture systemic inflammation and nutritional status. These included the neutrophil-to-lymphocyte ratio (NLR), prognostic nutritional index (PNI), AST/ALT ratio, BUN/creatinine ratio, and corrected calcium. The NLR was calculated as the ratio of absolute neutrophil to lymphocyte counts. The PNI was defined as albumin (g/dL) + 5 × lymphocyte count (10□/L), consistent with prior literature^61^. The AST/ALT and BUN/creatinine ratios were calculated directly from raw lab values, with thresholds for abnormality based on clinical references (e.g., AST/ALT > 2.0; BUN/Creatinine > 20). Corrected calcium was computed using the formula: calcium + 0.8 × (4 – albumin)^60^.

### Mixed Effects Regression Model

Mixed-effects logistic regression was performed to evaluate associations between cachexia status and clinical or biological variables in cancer patients. The models were fit using the glmer function from the lme4 R package, with a random intercept for each patient (MRN) to account for repeated measures. Models were stratified by cancer type and fit only in cancer types with ≥500 patients.

For serologic analysis, 30 laboratory markers were assessed. Each lab measurement was aligned to whether it occurred during a cachectic or non-cachectic period based on overlapping timestamps. The model included fixed effects for lab value, and sex. The following formula was used:

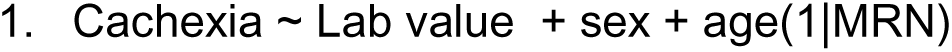

P-values were adjusted using the Benjamini-Hochberg correction.

### Permutation Analysis

To evaluate whether cachexia episodes occurred in temporal proximity to disease progression more frequently than expected by chance, we performed a permutation test for each cancer type. For each patient, the timing of cachexia episodes was randomly reassigned within their observed follow-up window while preserving the number, duration, and temporal structure of episodes. This process was repeated 1,000 times to generate a null distribution of expected overlap between cachexia episodes and radiographically defined progression events. The observed overlap in the original dataset was then compared to this null distribution to calculate empirical p-values, with significance defined as an observed overlap exceeding the 95th percentile of the permuted distribution.

### Weight-loss Grading System

To quantify cachexia severity, we applied the weight-loss grading system (WLGS) as described by Martin et al^25^. We extracted discrete cachexia episodes and used the BMI at episode start as the baseline. Episode-specific weight loss was then calculated relative to this baseline, and patients were classified into WLGS categories according to the matrix of percentage weight loss and baseline BMI.

### Survival Analysis

We evaluated the impact of cachexia and recovery on overall survival using Cox proportional hazards models. Both fixed and time-varying covariates were used to capture the onset and persistence of cachexia and recovery. In fixed models, patients were classified as cachectic if they experienced ≥1 cachexia episode. In time-varying models, cachexia status was updated at the onset of the first episode and remained active thereafter. Recovery was modeled similarly, beginning at the time of sustained BMI gain. All models were adjusted for age at diagnosis, sex, and cancer type using the coxph function from the survival R package.

The following equations were used.

1. Survival ∼ Cachexia + Age + Sex + Cancer Type + cluster (MRN)
2. Survival ∼ Cachexia (time varying) + cluster (MRN)
3. Survival ∼ Cachexia (time varying) + Age + Sex + Cancer Type + cluster (MRN)
4. Survival ∼ WLG (0-2) (time varying) + cluster (MRN)
5. Survival ∼ WLG (3-4) (time varying) + cluster (MRN)

### Time to cachexia analysis

To investigate the influence of somatic alterations on cachexia risk, we integrated mutation calls from the MSK-IMPACT panel with curated clinical covariates. For each cancer type with sufficient sample size (n > 200), we first identified candidate genes through univariate analyses (Benjamini–Hochberg adjusted *p* < 0.1). Only oncogenic mutations with a mutation frequency greater than 5% were tested. Cumulative incidence of cachexia in wildtype and mutant genes were calculated separately using the AalenJohansenFitter function in the *lifelines* Python package (v0.27.8). This method accounts for death as a competing event which might prevent the event of interest (cachexia) from ever occurring. Significant genes were then added into multivariate Cox proportional hazards models, where the outcome was time from tumor diagnosis to cachexia onset with death treated as a competing event. Models were stratified by cancer type and adjusted for demographic and clinical features, including age at diagnosis, sex, sample type, TMB, disease stage, ECOG performance status, and baseline BMI (categorized and scaled as described). For colorectal, gastric, and endometrial cancers, MSI status was additionally included, and for colorectal cancer we also adjusted for tumor sidedness. Models were fit using the CoxPHFitter function in the *lifelines* Python package (v0.27.8), with the following equation:

1. Univariate: time_to_cachexia ∼ gene
2. Multivariate: time_to_cachexia ∼ sig_gene + age+ sex + starting BMI + stage + TMB + ECOG + sample type
3. Multivariate (gastric, endometrial): time_to_cachexia ∼ sig_gene + age+ sex + starting BMI + stage + TMB + ECOG + sample type + MSI
4. Multivariate (colorectal): time_to_cachexia ∼ sig_gene + age+ sex + starting BMI + stage + TMB + ECOG + sample type + MSI + sidedness

### Nomogram construction and calibration

To provide individualized prediction of cachexia risk, we constructed nomograms based on multivariable Cox regression models using the covariate framework that were tested significantly associated with cumulative incidence of cachexia. Models were trained on cancer-specific cohorts, and the regression coefficients were scaled to assign point values to each predictor. The cumulative point total was mapped to the predicted probability of cachexia at fixed time horizons (1-, 2-, and 3-years post-diagnosis).

The predictive performance of the models was assessed for discrimination and calibration using an internal validation approach. The full cohort for each cancer type was randomly split into a training set (50% of patients) and a testing set (50% of patients). Discrimination, the model’s ability to distinguish between patients who develop cachexia early versus late, was evaluated using two metrics: (1) We performed bootstrapping with 200 resamples to calculate the mean concordance index and its corresponding 95% CI. (2) Dynamic Time-Dependent Area Under the Curve (AUC) were calculated for 1-, 2-, and 3-year cachexia risk prediction using the cumulative_dynamic_auc function from the Python *sksurv* package.

We measured calibration, the agreement between the model’s predicted probabilities and the actual observed outcomes by generating calibration plots for the test cohort. Patients in the test set were stratified into quartiles based on their nomogram-predicted risk of cachexia. Within each quartile, the mean predicted risk was plotted against the observed cachexia incidence. A perfect calibration would present a 45-degree diagonal line.

## Data Availability

Data is available in **Supplementary Tables**. Code for reproducing the results and associated tables is available at https://github.com/reznik-lab/cachexia.

## Acknowledgements

We thank the Reznik, Janowitz, and Goncalves labs for useful feedback and Dr. Christina Campagna for inspiring and thought-provoking daily discussions. This work was supported by NCI P30 CA008748. E.R. was supported by NIH R37 CA276200, DOD HT9425-23-1-0995, a Kidney Cancer Association Young Investigator Award, the MSK Society, and the Pershing Square Sohn Cancer Research Foundation. This work was also supported by Cancer Grand Challenges CGCSDF-2021\100003 (CRUK) and 1OT2CA278609-01 (NCI).

---

**Supplemental Figure 1.**
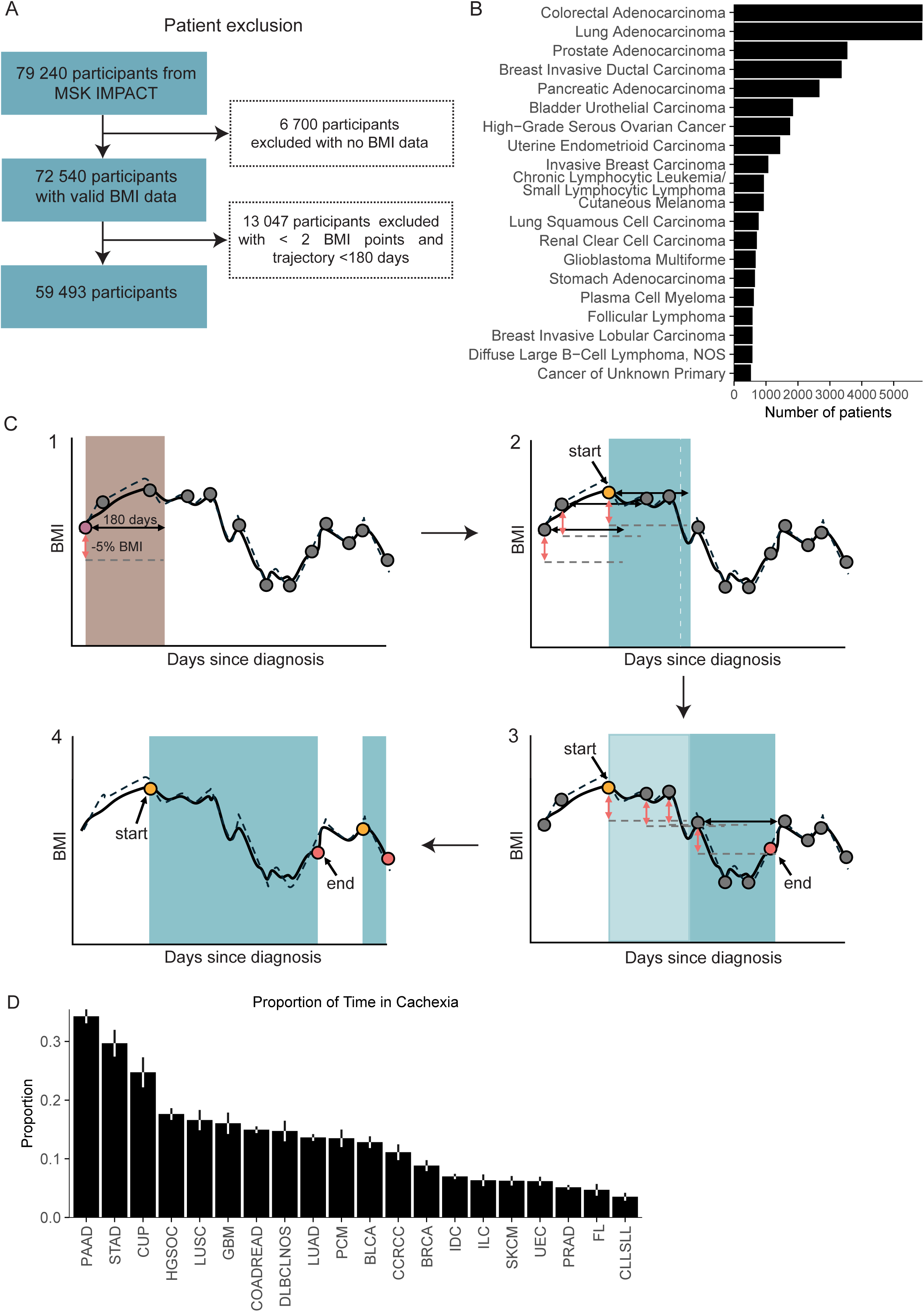
Identification of cachexia episodes. (**A**) Flowchart of patient exclusion. (**B**) Cancer type distribution. **(C)** Schematic illustrating the sliding-window approach used to define cachexia episodes (≥5% weight loss over 180 days), merging overlapping windows to capture sustained weight loss. (**D**) Proportion of total clinical follow-up spent in cachexia, stratified by cancer type.

**Supplemental Figure 2.**
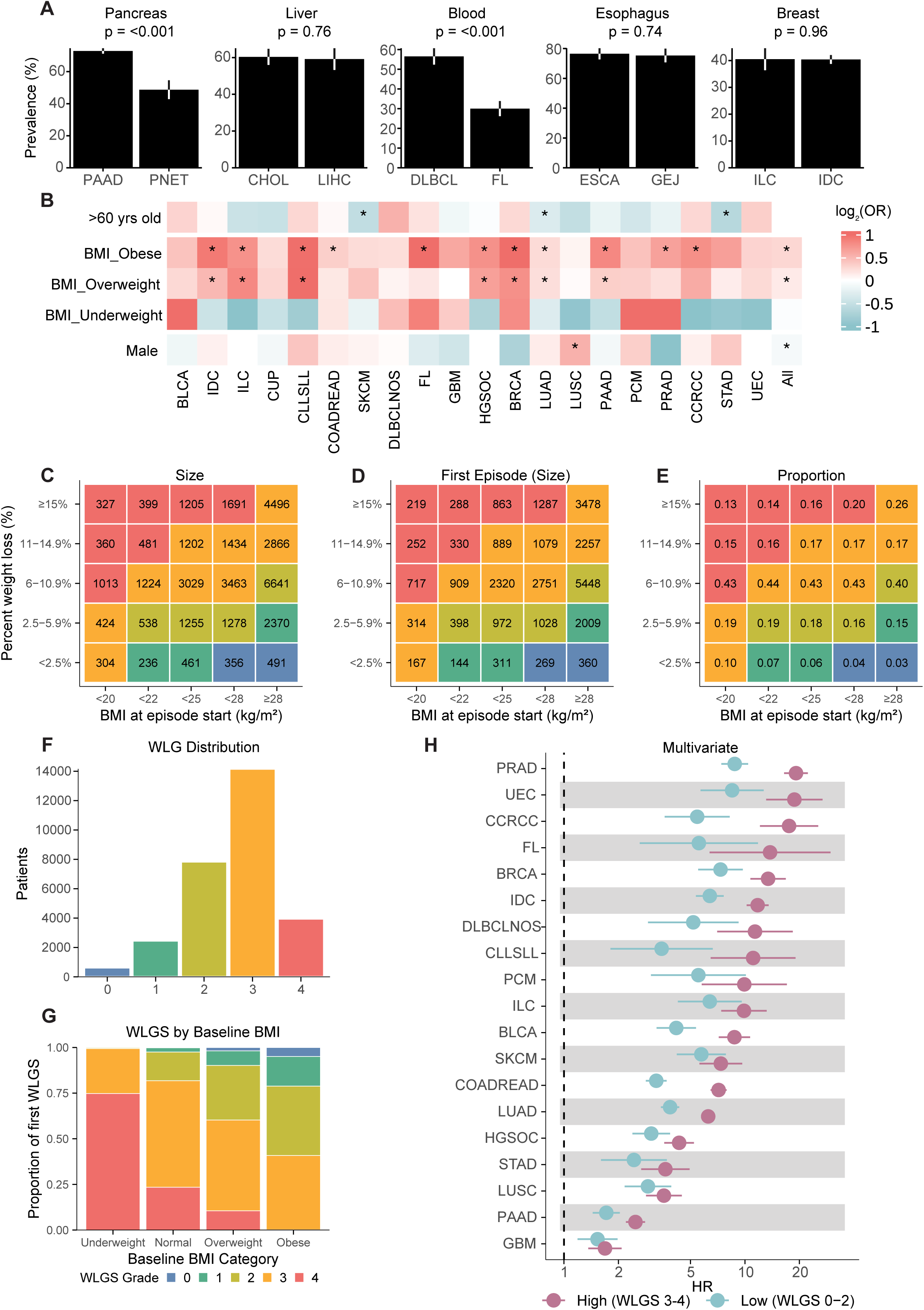
**(A)** Logistic regression showing associations of sex, age, and BMI at diagnosis with cachexia across cancer types **(B)** Cachexia prevalence within selected tissues of origin: pancreas (PAAD vs. PNET), liver (CHOL vs. LIHC), blood (DLBCL vs. FL), esophagus (ESCA vs. GEJ), and breast (ILC vs. IDC). **(C)** WLG (0–4) size matrix of discrete cachectic episodes by BMI and weight loss **(D)** WLG (0–4) size matrix of first cachectic episodes by BMI and weight loss **(E)** WLG proportion matrix of first cachectic episode incidence by BMI category. **(F)** Distribution of WLG groups across the cohort. **(G)** Proportion of WLG distribution by baseline BMI category **(H)** Time-dependent hazards for overall survival between non-cachectic patients with WLG 0–2 (low) and WLG 3–4 (high).

**Supplemental Figure 3.**
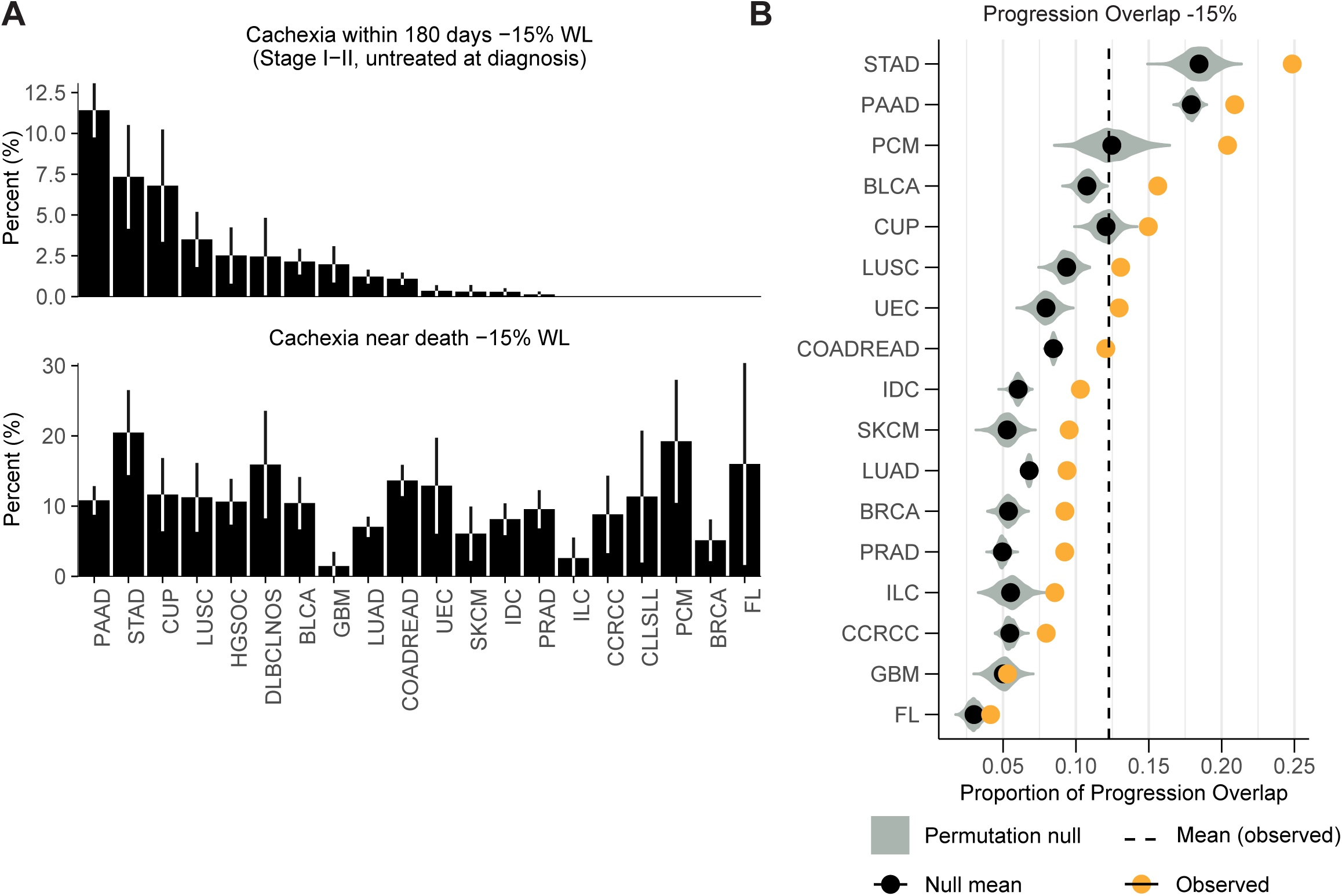
**(A)** Rates of cachexia within the first 180 days of diagnosis (Stage□I– II, untreated at diagnosis; top) and near-death (bottom) for ≥15% threshold. **(B)** Summary of cachexia–progression overlap across cancer types using ≥15% WL threshold.

**Supplemental Figure 4.**
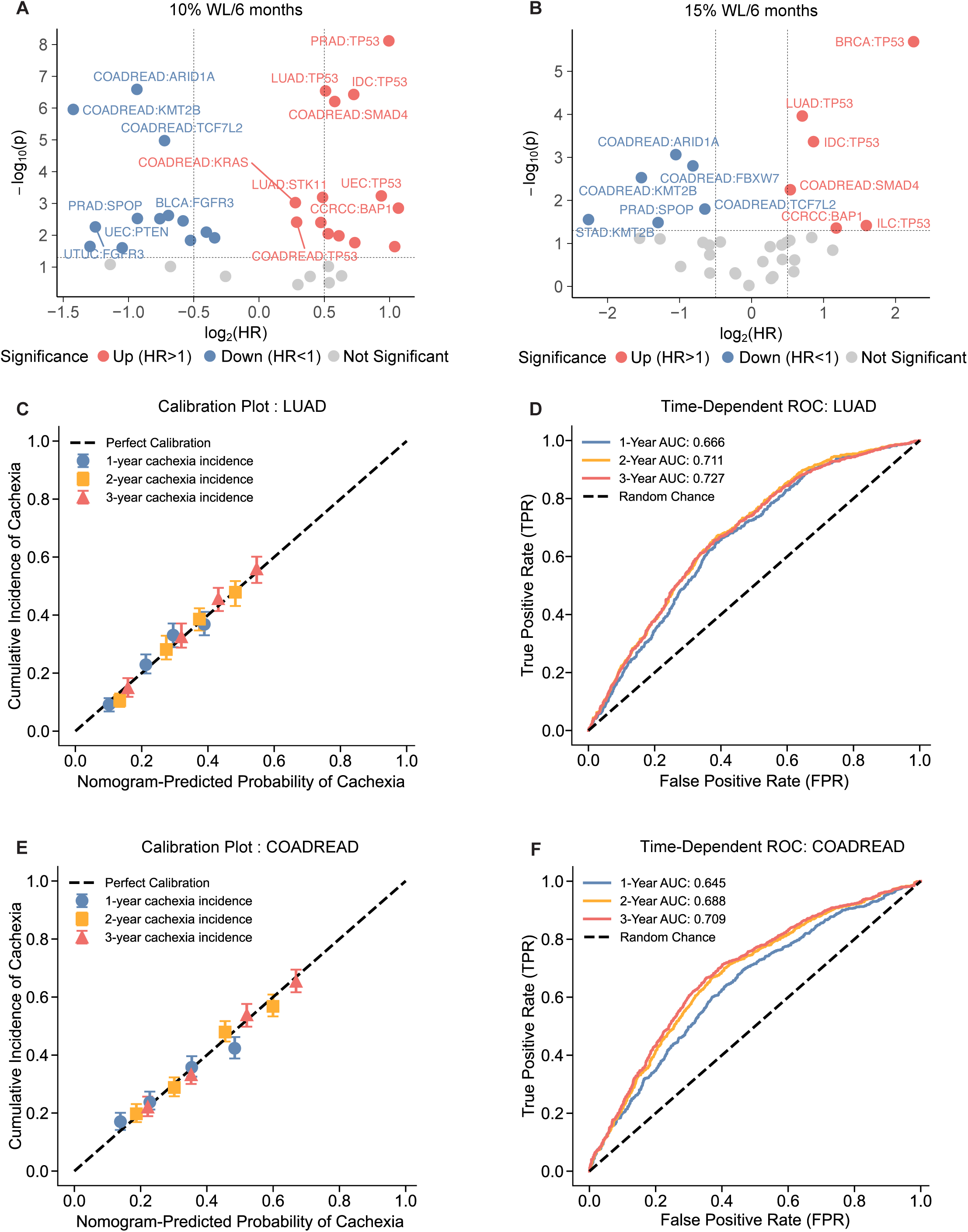
**(A)** Univariate association between the presence of driver mutations and risk of cachexia from a competing risks Cox regression using a ≥10% over 6-month period weight loss threshold to call cachectic episodes. **(B)** Univariate association between the presence of driver mutations and risk of cachexia from a competing risks Cox regression using a ≥15% over 6-month period weight loss threshold to call cachectic episodes. **(C–F)** Performance of cachexia risk models. Calibration **(C, E)** of predicted versus observed cachexia incidence at 1, 2, and 3 years in lung adenocarcinoma and colorectal adenocarcinoma, respectively. Time-dependent ROC curves **(D, F)** for cachexia risk prediction, with AUCs of 0.666, 0.711, and 0.727 for lung adenocarcinoma and 0.645, 0.688,0.709 for colorectal adenocarcinoma at 1, 2, and 3 years.

**Supplemental Table 1:** Univariate Logistic Regression Results Across Cancer Type (Demographics)

**Supplemental Table 2:** Multivariate Time-Dependent Cox Model Results Across Cancer Type

**Supplemental Table 3:** Relative odds of early cachexia onset across tumor types (5%,10%,15%)

**Supplemental Table 4:** Mixed-effects Models Results Across Cancer Type-Lab pairs

**Supplemental Table 5:** Univariate Competing-Risks Regression Results Across Cancer Type-Gene Pairs for 10% threshold/6 months (p<0.05).

**Supplemental Table 6:** Univariate Competing-Risks Regression Results Across Cancer Type-Gene Pairs for 15% threshold/6 months (p<0.05).

**Supplemental Table 7:** Clonal Validation of Genotype-Cachexia Associations by Univariate Competing-Risks Regression (5% WL/6 months)

**Supplemental Table 8:** Multivariate competing-risks regression for cachexia in LUAD (FDR<0.05)

**Supplemental Table 9:** Multivariate competing-risks regression for cachexia in COADREAD (FDR<0.05)

**Supplemental Table 10:** Cancer patient summary

